# Reward processing in young people with self-harm behaviour

**DOI:** 10.1101/2023.12.16.23300091

**Authors:** Emre Yavuz, Rachel Rodrigues, Ana Pascual Sanchez, Anne Lingford-Hughes, Martina Di Simplicio

## Abstract

Twenty percent of young people report a lifetime presence of self-harm (SH) behaviour, associated with negative health and functional outcomes. Understanding the underlying cognitive mechanisms is needed to develop targeted early interventions. Reward processing biases may underlie SH, aligning with accounts of the behaviour acquiring “addictive” characteristics. However, the specific nature of such biases remains unclear, particularly its relationship with negative affect (NA) that frequently triggers SH. In Study 1, we compared young people (aged 16-25) with SH to a group with NA but no SH history and a healthy control group on performance of a novel Incentive Delay Task (IDT), with SH-related (SH trials), positive social (social trials) or monetary images (money trials) as stimuli. In Study 2, a different sample of SH and HC participants completed the same IDT following NA induction via an online Trier Social Stress Test. For both studies, we hypothesised faster and more correct responses in the SH group than control groups on SH trials. Contradicting our hypothesis, there were no significant between-group differences in IDT performance on SH, social and money trials in either study. Certain SH characteristics (positive reinforcement, SH mental imagery, urge) were significantly correlated with better performance on SH trials in SH participants. Thus, broadly SH behaviour may not be underpinned by motivational biases towards SH-related cues or naturalistic rewards. Future studies should clarify whether incentivisation of SH-related cues instead explains individual differences in SH behaviour and its relation with treatment and prognosis.

## Introduction

Self-harm (SH) is defined as “any act of self-injury or self-poisoning carried out by a person, irrespective of their motivation”^1^. SH most commonly occurs in young people by self-cutting, occurring in about 18% of adolescents^2–7^. Engaging in SH increases risk of future suicide attempts and other adverse outcomes including substance abuse and impaired psychosocial functioning^3^, thus early intervention is important.

SH is understood as a manifestation of emotional distress and is often used to cope with difficult emotions^2^. As the severity and frequency of SH varies widely^2–7^, it is essential to understand the motivational processes that underlie self-harm and may differentiate those that self-harm repeteadly. In keeping with personal accounts of self-harm^7^, this has also been conceptualised as an addictive behaviour that can be learned and repeated as a result of reinforcement from reducing negative and/or enhancing positive emotions^4–6^. Hence, an understanding of the underlying mechanisms of this addictive component could aid the development of preventative strategies for repeated SH.

One of the theoretical frameworks for understanding addictive behaviours is the Incentive Sensitisation Theory^8^. Incentive sensitisation is thought to occur due to reward system hypersensitisation to a substance or behaviour and its associated cues, and helps explain how the occasional use of substances can lead to addiction or relapse^8^. According to this theory, substance-related cues become conditioned and predictive of reward, resulting in attentional resources being preferentially allocated to them^8^. Exposure to these cues can elicit reward anticipation and motivation to obtain the reward^9^. Initial evidence supports the idea that SH-related cues may also become sensitised and elicit reward anticipation^10,11^. These SH cues may include external cues such as objects to self-harm or internal representations, such as mental imagery of SH^12^, which is highly prevalent in those who SH^13^ and could play a role similar to mental imagery of substances, a key feature of craving in addiction^14^.

Past research suggests these biases in reward processing systems may be present in SH^15–17^. For example, a genetic predisposition to lower dopamine transmission is correlated with greater SH in individuals with bulimia^15^ and in individuals who experienced child abuse^16^. Using fMRI, neural activity in reward-related regions has been shown to be both higher^18.19^ and lower^17^ when anticipating monetary rewards in SH groups compared with healthy controls, which may be associated with predisposition to SH like in other addictive behaviours^20^. However, studies also report no differences in sensitivity to social rewards in SH groups compared with controls^21,22^.

The term ‘waiting impulsivity’ has also been used to characterise other addictive behaviours aside SH, describing the ability to refrain from responding until signalled to do so by explicit cues^23–25^. These inconsistent findings highlight the need to clarify the presence and direction of motivational biases for natural rewards in SH. Given that greater self-reported impulsivity has also been associated with a greater likelihood of SH behaviour^23^, this could be explored in the context of different reward stimuli.

Past research also suggests that individuals engage in SH to relieve negative affect (NA)^6^. Greater NA often preceeds SH enactment^26,27^ In line with this, NA has been reported in individuals with SH compared with healthy controls in cross-sectional and longitudinal behavioural studies^28–30^. In young people with SH, BOLD activity in reward-related regions correlates with self-reported deficits in emotion regulation^31^, and is modulated by stimuli’s valence^32^. NA is also associated with reduced reward signalling for non-salient rewards in addiction, often leading to relapse and increasing motivation for the addictive behaviour^33^. For example, some young people frequently report that they self-harm because that provides more intense stimulation than other alternative strategies. However, the interaction between NA and reward processing remains unexplored in those with SH.

We conducted two studies to investigate motivational biases for SH-related stimuli and natural rewards in young people with SH (Study 1). In a separate sample of participants, we investigated how these biases are influenced by NA using a NA induction (Study 2). We used a novel behavioural Incentive Delay Task (IDT^34^) with money, positive social and SH cues, measuring the number of correct responses and reaction latency across trials (motivational bias). We also measured the number of premature responses across trials as an indicator of waiting impulsivity.

In Study 1 we compared individuals with SH to both a HC group, and a group with equivalent levels of NA but no SH history, to control for the effect of low mood and anxiety on reward processing and explore why some individuals do not SH despite experiencing high NA^2,35^. As motivational biases may only be elicited in the presence of NA, a different SH and HC group repeated the same experiment in Study 2 after undergoing the Trier Social Stress Test. Our primary hypothesis for both studies was that if SH participants were biased to salient SH-related stimuli, the SH group would respond faster to-and make more correct or premature responses to SH stimuli compared with NA and HC groups in the IDT. As findings on motivational biases for natural stimuli in SH are inconsistent, we explored differences in RT, accuracy and premature responses to social or monetary rewards in the IDT. We also explored the association between responses to SH stimuli and self-reported measures of SH frequency, duration and recency, endorsed motivation for SH, urge to SH, SH imagery characteristics (Study 1 only) in the SH group, and state depression, anxiety and stress levels in SH and NA groups.

## Methods

We collaborated with a Young Person’s Research Group (YPRG) formed of young people aged 16 - 25 with lived experience of self-harm and other mental health difficulties to deliver these studies. The YPRG contributed to the experimental design, development of the IDT, recruitment methods and interpretation of results. The study was approved by the NRES Committee South Central Oxford-C (REF: 19/SC/0275). All materials and data are reported on Open Science Framework (https://osf.io/8kcg3/).

### Participants

Participants were recruited using social media advertisements on Instagram and Facebook (see Supplementary Material for details) and reimbursed with a £50 voucher for completing the testing. Inclusion criteria for all 3 groups included being 16-25 years old, having adequate English to provide informed consent and understand the study and the tasks. Exclusion criteria included having severe neurological impairment, learning disability, or current, acute psychotic symptoms that could interfere with task performance. The group-specific inclusion criteria were: 1) SH group having at least one episode of SH within the past year; 2) NA group having depression and/or anxiety scores on the Depression, Anxiety and Stress Scale (DASS-21)^36^ above the normal range (depression score >= 10, anxiety score >= 8, stress score >= 15); healthy controls (HC) group having DASS-21 depression, anxiety and stress scores within the normal range. Participants were excluded from the NA group if they had a history of SH or suicide, current or past substance abuse or dependence, as well as if they had smoked regularly or vaped nicotine (self-reported), assessed using the MINI International Neuropsychiatric Interview (MINI)^37^ and smoking assessment. Participants were excluded from the HC group if they had presented a history of mental health difficulties, SH, suicide or substance misuse or dependence, as well as if they had smoked regularly or vaped nicotine (self-reported), also assessed using the MINI and smoking assessment.

### Experimental Procedure

The testing session for studies 1 and 2 was conducted online over MS Teams by members of the Mood Instability Research Group. Both studies took place online given the COVID pandemic, with study 1 taking place from January 2019-2021 and study 2 from January 2021-2023.

#### Self-report questionnaires

Participants answered self-report questionnaires via Qualtrics links shared by the experimenter over MS Teams. Clinical characteristics were assessed via the MINI^37^ and the McLean Screening Instrument for Borderline Personality Disorder (MSI-BPD)^38^. Questionnaires relating to SH included the Self-Injurious Thoughts and Behaviour Interview (SITBI)^39^, the Self-Harm Imagery Interview (SHII)^40^ (Study 1 only), the Alexian Brothers Urge to Self-Injure Scale (ABUSI)^41^ and the Craving Experiences Questionnaire adapted for SH (CEQ-SH)^14^ (Study 2 only). State depression, anxiety and stress was assessed using the DASS-21^42^. The Positive and Negative Affect Schedule (PANAS) was administered before and after the Trier Social Stress Test in Study 2 (see below). See Questionnaires details in the Supplementary Materials.

#### Incentive Delay Task

This study used a novel IDT developed by our team and adapted from the original fMRI Monetary Incentive Delay paradigm^17,34^, Participants completed the task on the online platform Pavlovia. The task had 3 reward types: SH, positive social and money, with two blocks per reward type and 50-60 trials/block. For each reward type, half the trials were a neutral condition and half were a ‘win’ condition with reward-based stimuli (SH, positive social, money). For the SH reward type, half the SH-related stimuli were standardised across tasks and half were tailored to each participant’s SH methods. NA and HC participants tasks were matched to SH participants. The order of the reward types was counterbalanced across participants.

During the task, a cue appeared which predicted the type of feedback image (reward) participants would receive, followed by a target (black square) which then disappeared. Participants had to respond correctly by pressing a key before the target disappeared to obtain the feedback images. If the target was correctly responded to, a neutral image (either an image of a neutral object, a neutral social scene or an image of an empty container; neutral condition) or reward image (either an image of act of SH, a positive social scene or an image of money in a container; win condition) appeared (Figure 1). See Supplementary Figure S1 for details of the IDT stimuli. Participants also won 20p per correct response in win money trials, as informed in the pre-task instructions. If participants responded to the cue, the feedback ‘Too early’ appeared. If they responded after the target disappeared, the feedback ‘Try to be faster’ appeared. The target’s minimum and maximum duration of presentation was calculated based on the standard deviation of the participant’s baseline RTs from the fastest half of trials during a 20-trial practice block (based on a previous study^43^). This ensured sufficient task difficulty.

**Figure 1:**
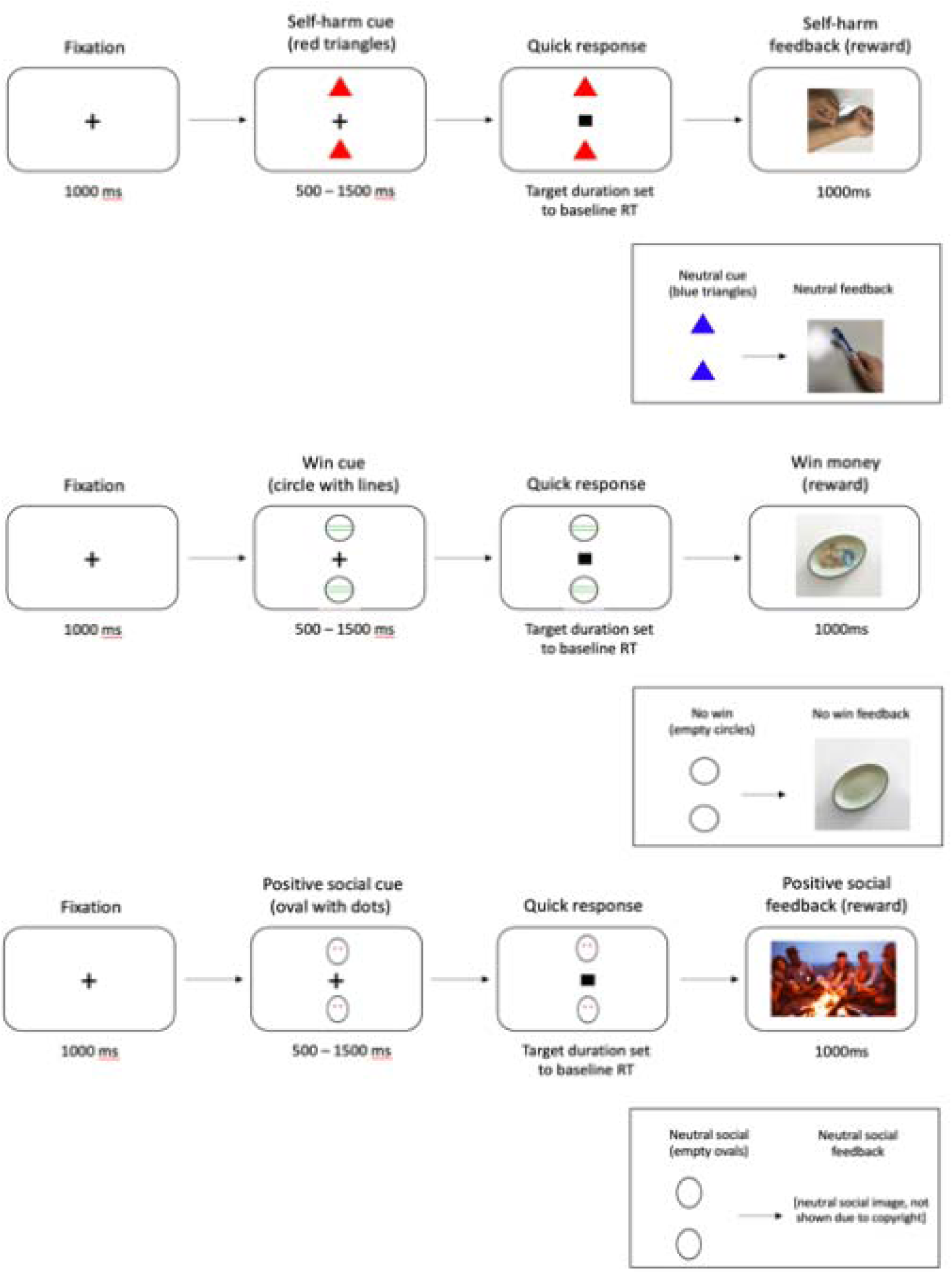
Incentive Delay Task, showing the self-harm reward type (top), money reward type (middle) and social reward type (bottom).

Accuracy and mean RTs on the IDT were calculated for correct responses to the target. Accuracy was calculated by summing the number of correct target responses occurring after 0.1s from the target’s appearance for reward and neutral conditions, for each reward type separately (log transformed). Premature responses, measuring waiting impulsivity, were calculated by summing the number of responses to the cue and target that occurred within 0.1s of the target’s appearance for reward and neutral conditions, for each reward type separately (log-transformed).

In Study 1, after IDT completion, participants also rated each of the SH stimuli from 0 (not at all) to 10 (extremely) on five scales: calming, exciting, pleasant, unpleasant and distressing, creating one score for each scale. These 5 scores were also summed to create a total score given previous findings suggesting that SH elicits mixed emotions^44,45^. These scores were used to explore whether perception of SH stimuli influences behavioural responses during the IDT.

#### Trier Social Stress Test

In Study 2, negative affect was induced by administering the Trier Social Stress Test (TSST). The TSST is the most frequently used social evaluative stress task in children, adolescents and adults^46,47^, and effectively induces self-reported NA and physiological stress responses in HC and SH groups^48,49^. An online TSST^50^ was used based on the original in-person TSST^51^ and shown to be effective in inducing self-reported and physiological stress responses in healthy adolescents^50^.The TSST has 3 sections: a 5-minute speech preparation period; a 5-minute speech presentation; and a 5-minute arithmetic task. The speech and arithmetic task are completed in front of 2 judges (Figure 2). See Supplementary Materials for details.

**Figure 2.**
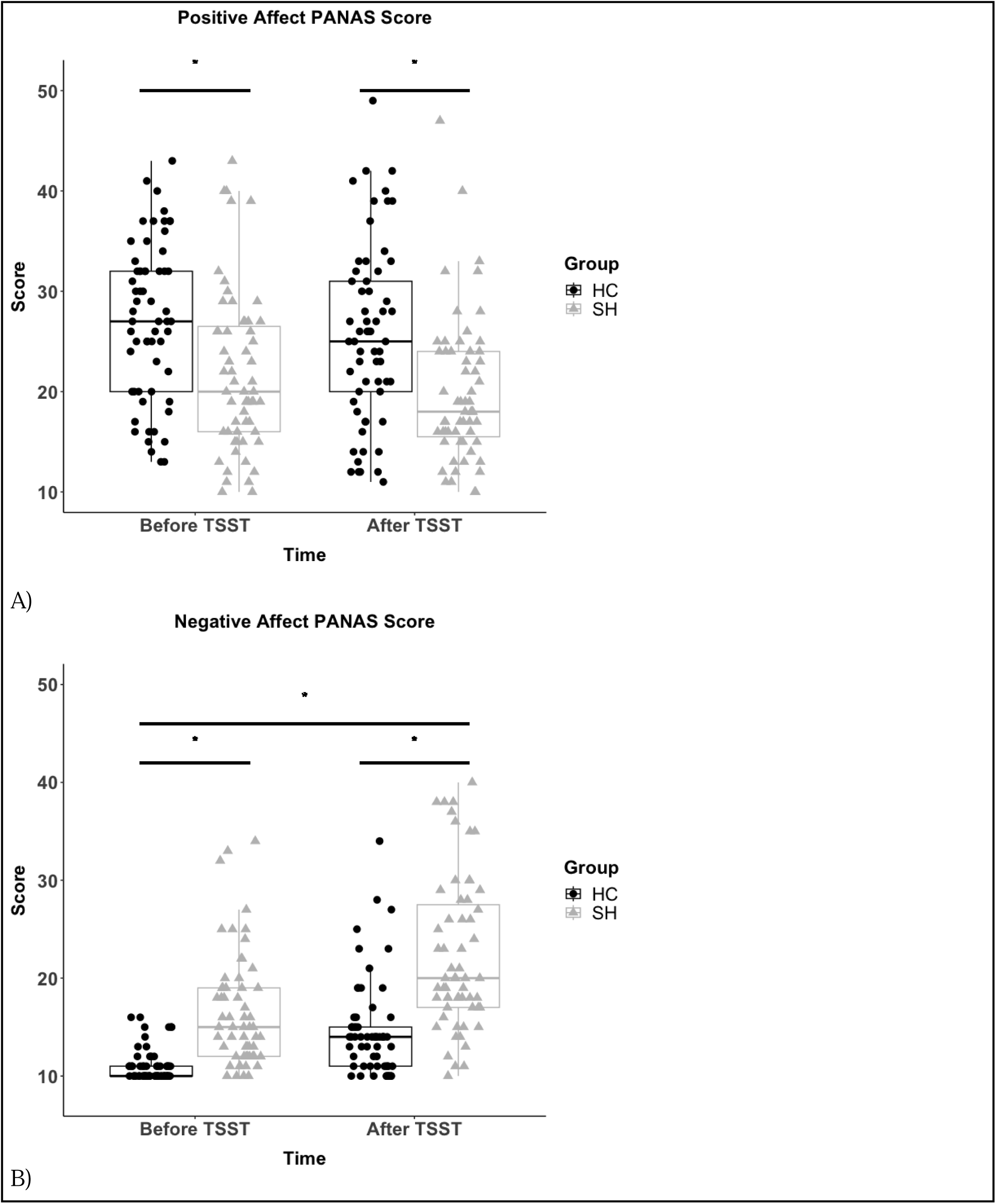
Change in Positive Affect PANAS scores (A) and Negative Affect PANAS scores (B) from before to after the TSST in Study 2. * = p < 0.05.

### Statistical analysis

A power analysis (based on an Incentive Delay Task used by a previous study comparing reaction times in SH compared HC participants^17^) indicated that N=48 participants were required per group for an effect size of d = 0.580 at 80% power, with an alpha threshold of 0.05 (see Supplementary Materials for details).

In Study 1, correct and premature responses were analysed using a 3 x 3 x 2 ANOVA, with group (SH, NA and HC) as a between-group factor and reward (SH, social and money) and condition (win, neutral) as within-group factors. Significant main effects of group and reward were examined using Bonferroni-corrected post-hoc t-tests or Games-Howell tests where homoscedasticity was not met, and significant interactions were analysed using simple main effects analyses. Total money won was compared between groups using a one-way ANOVA, with significant main effects of group examined with Bonferroni-corrected post-hoc t-tests. A reward-neutral trials contrast was calculated for the percentage of correct trials, premature responses and RTs for correct responses in the IDT to run correlational analysis. Each of these 3 contrasts were correlated with past-year, lifetime frequency and duration of SH and endorsement of automatic positive or negative reinforcement as SH motivator (SITBI); urge to SH (ABUSI and CEQ); characteristics of self-harm-related mental imagery (Self-Harm Images Interview)(Study 1 only); and state depression, anxiety and stress (DASS-21) scores using Spearman-rank correlations. As these correlations were exploratory, we did not correct for multiple comparisons.

The analysis procedure for the IDT in Study 2 was identical to part 1, except only 2 groups (SH vs HCs) were used for analysis and SH-related imagery scores were not available, as mentioned above. Additionally, two separate contrasts for positive and negative PANAS scores respectively (PANAS scores after - before TSST) were correlated with contrasts for each of the IDT outcome measures (reward - neutral trials) for each group and for each condition separately using Spearman-rank correlations. This was to assess whether TSST-induced changes in negative or positive affect were associated with a motivational bias in the IDT.

## Results

### Study 1

Fifty-four SH, 56 NA and 51 HC participants initially completed the IDT task. After removal of outliers (see Supplementary Materials), a final sample of 49 SH, 54 NA and 49 HC was left. Demographic and SH characteristics are reported in Table 1 and 3 respectively. Clinical characteristics are reported in Supplementary Table S4.

**Table 1.** Self-harm characteristics of SH group in Study 1 and 2. ABUSI: Alexian Brothers Urge to Self-harm scale.

|  | Study 1 |  | Study 2 |  |
| --- | --- | --- | --- | --- |
| Self-harm Characteristic | Mean (± SD) | Median (IQR) | Mean (± SD) | Median (IQR) |
| **Recency:** No. days since last self-harm episode | 72.6 (± 90.7) | 30 (143) | | |
| **Lifetime frequency** | 396.5 (± 829.5) | 100 (217) | 283.6 | 46.5 (234.5) |
| **Past-year frequency** | 52.6 (± 829.5) | 100 (217) | 41.2 (± 72.0) | 10 (31.5) |
| **Past-month frequency** | 3.2 (± 6.0) | 1 (4) | | |
| **Age onset, yrs** | 13.8 (± 3.5) | 13 (4) | 13.5 (± 2.4) | 13 (3) |
| **Duration, yrs** | 5.5 (± 3.6) | 5 (5) | 4.8 (± 3.2) | 4 (3) |
| **Number of self-harm methods** | 2.4 (± 1.2) | 2 (1) | 1.5 (± 0.8) | 1 (1) |
| **Ever needed medical treatment** (yes:no:unknown) | 15:39:0 | - | 15:61:4 | - |
| **Future likelihood of self-harm** (0-4 scale) | 2.7 (± 1.2) | | | |
| **Positive reinforcement endorsement score** | 2.33 (± 1.2) | 3(2) | 2.39 (± 1.3) | 3(1) |
| **Negative reinforcement endorsement score** | 2.67 (±0.9) | 3(1) | 2.95 (± 0.9) | 3(2) |
| **Urge to self-harm** (ABUSI, max score 35) | 16.7 (± 6.8) | 15 (10) | 12.0 (± 7.2) | 12 (11.5) |

#### Incentive Delay Task performance

There was a significant main effect of group on correct responses (F(2, 148) = 4.98, *p* = 0.008, *ηp2* = 0.06), where post-hoc t tests showed that the SH group made more correct responses to the target across the entire task than the NA group (p = 0.006) but not HCs (*p* = 0.215) (Figure 3). This significant difference was present for all 3 reward types (group x reward type x condition interaction: (F(4, 296) = 2.37, *p* = 0.053, *ηp2* = 0.03). There was a significant main effect of reward type for correct responses (F(2, 296) = 44.33, *p* < 0.001, *ηp2* = 0.23), where all participants had fewer correct responses in win and neutral money trials than in positive and neutral social trials and SH and neutral SH trials (*p* < 0.001). There was no significant main effect of condition (win vs neutral) for correct responses across all reward types (F(1,148) = 0.16, *p* = 0.691, *ηp2* = <0.01). However, there was a significant reward type x condition interaction (F(2, 296) = 6.76, *p* = 0.001, *ηp2* = 0.04), with a higher number of correct responses in the win money trials compared with the neutral money trials (*p* = 0.001), but not in SH ‘win’ trials compared with neutral SH trials (*p* = 0.423) and ‘win’ (positive) social trials compared with neutral social trials (*p* = 0.072).

**Figure 3.**
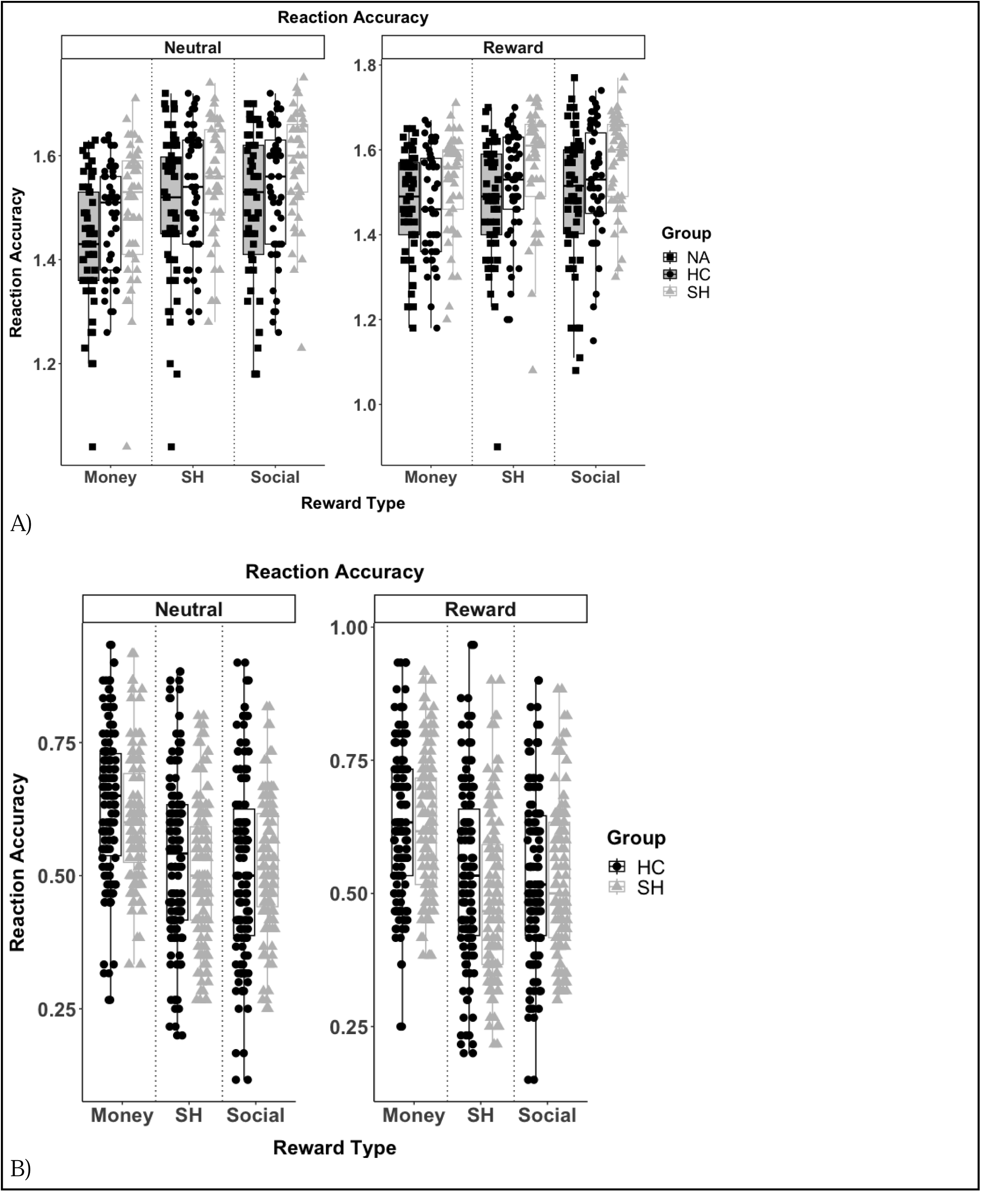
Correct responses for the Incentive Delay Task in Study 1 (A) and Study 2 (B) of the study.

There was no significant main effect of group on correct RTs to the target (F(2, 149) = 1.63, *p* = 0.199, *ηp2* = 0.02)(Figure 4). There was a significant main effect of reward type for correct RTs (F(2, 298) = 5.49, *p* = 0.005, *ηp2* = 0.04), with significantly shorter RTs for the money reward type than the SH reward type (*p* < 0.05). There was a significant main effect of condition (F(2, 149) = 23.79, *p* < 0.001, *ηp2* = 0.14), with significantly shorter correct RTs in win trials compared with neutral trials across all reward types (*p* < 0.005). There was a significant reward type x condition interaction for correct RTs (F(2, 298) = 16.91, *p* <0.001), with significantly shorter RTs in win money compared with neutral money trials (*p* < 0.001), but not in SH ‘win’ trials compared with neutral SH trials (*p* = 0.78) and ‘win’ (positive) social compared with neutral social trials (*p* = 0.248). There was no significant group x reward type x condition interaction for correct RTs (*p* > 0.05).

**Figure 4.**
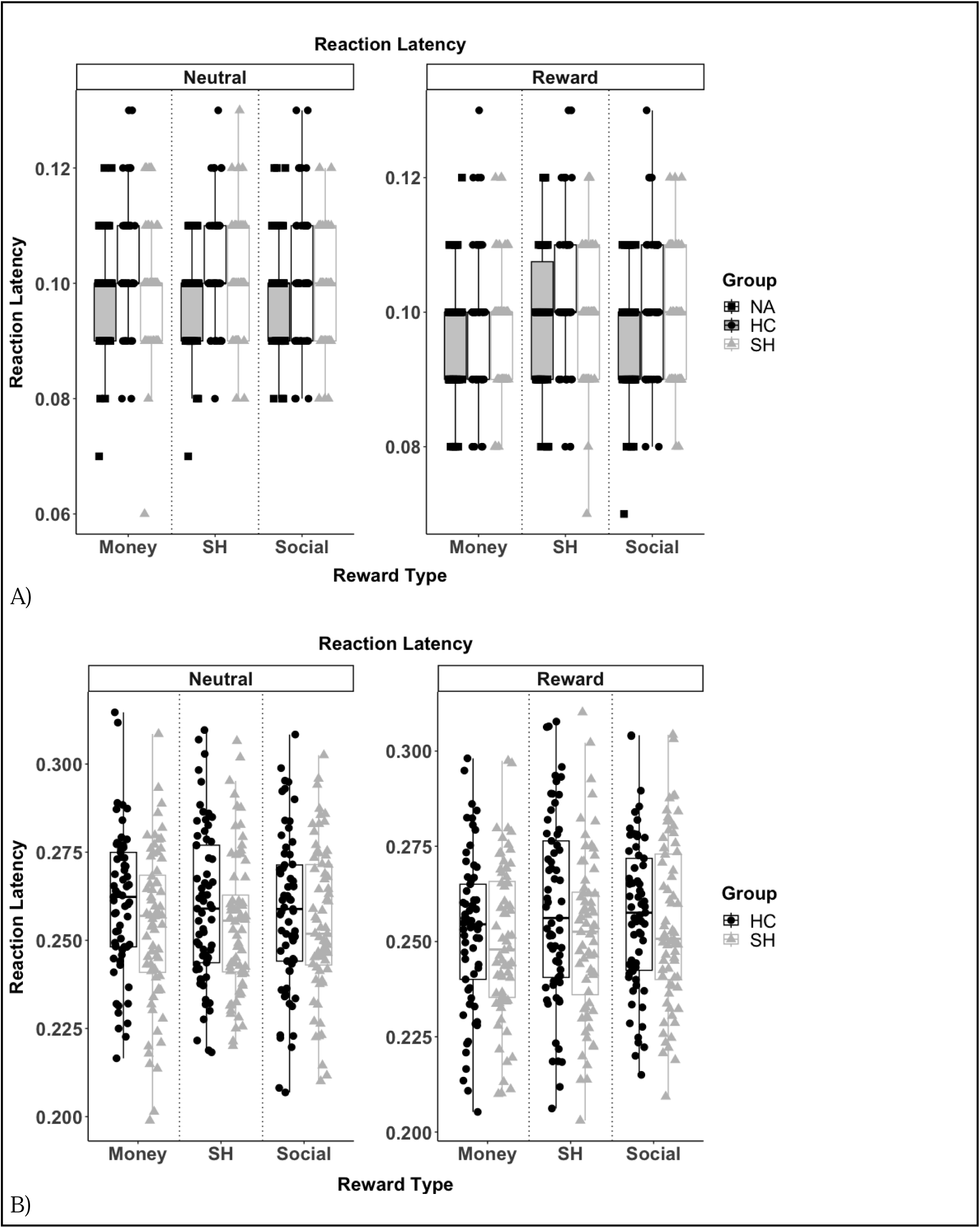
Reaction latency for the Incentive Delay Task for Study 1 (A) and Study 2 (B).

There was a significant main effect of group for premature responses (F(2, 147) = 3.57, *p* = 0.031, *ηp2* = 0.05), although between-group differences did not reach significance (*p* > 0.05)(Figure 5). There was a significant main effect of reward type (F(2, 294) = 23.25, *p* < 0.001, *ηp2* = 0.14), where all groups made significantly fewer premature responses for the money reward type than the SH and social rewards (*p* < 0.001). There was a significant reward type x condition interaction (F(2, 294)) = 19.46, *p* < 0.001, *ηp2* = 0.12), with all groups making more premature responses on SH ‘win’ trials relative to neutral SH trials and on win money trials relative to neutral money trials (*p* < 0.001), but not on ‘win’ (positive) social compared with neutral social trials (p=0.841). There was no significant group x reward type x condition interaction for premature responses (F(4, 294) = 1.42, *p* = 0.229, *ηp2* = 0.02).

**Figure 5.**
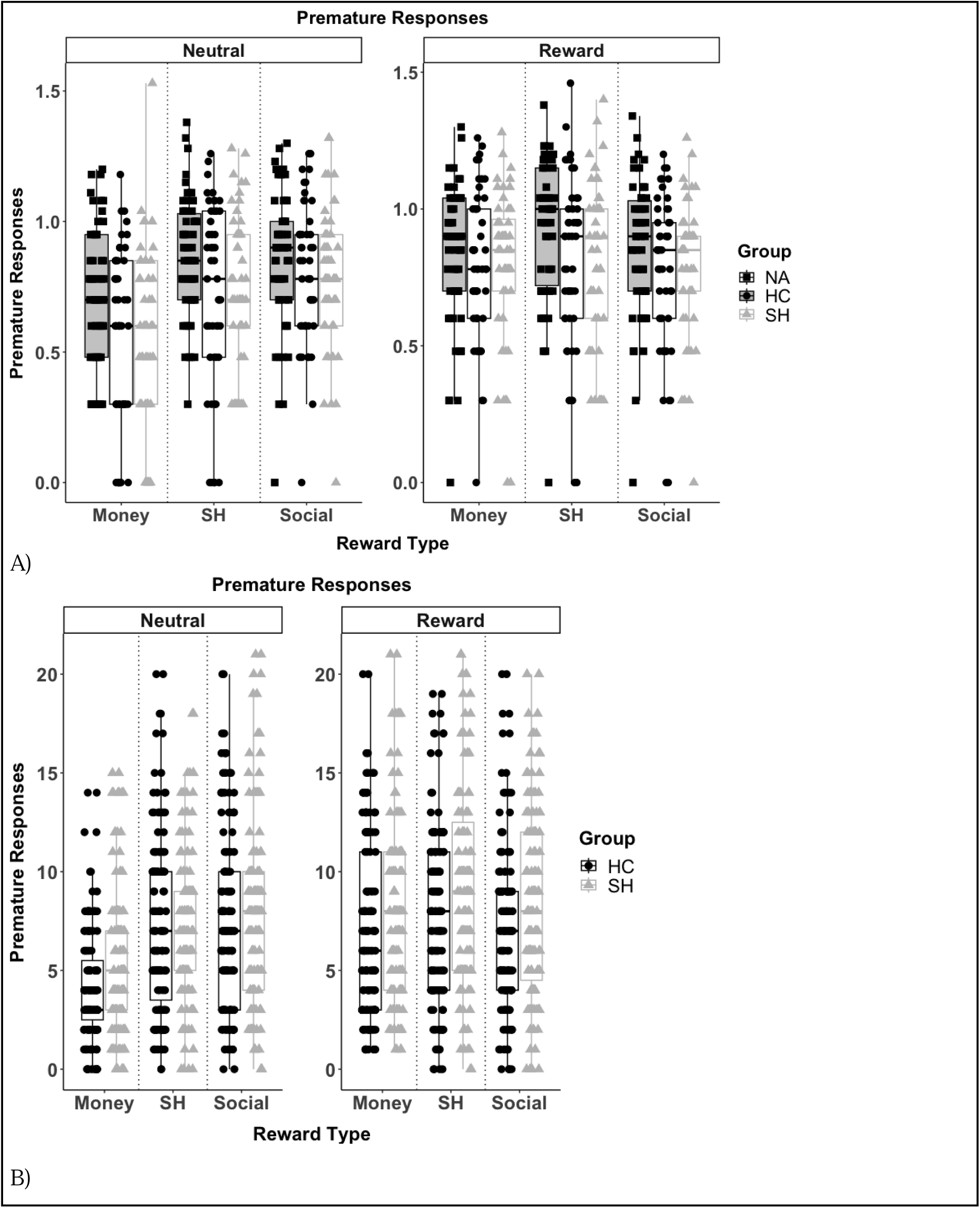
Premature responses for the Incentive Delay Task for Study 1 (A) and Study 2 (B).

There was a significant main effect of group for money won (F(2, 149) = 10.03, *p* = 0.025, *ηp2* = 0.05), with the SH group winning significantly more money than the HC group (*p* = 0.041), but not the NA group (*p* = 0.073)(Supplementary materials).

#### Associations between Incentive Delay Task performance and self-report questionnaires

Participants in the SH group who had been self-harming for fewer years had significantly more correct responses on SH trials (*rs* = 0.33, *p* = 0.024). Participants in the SH group who reported greater automatic positive reinforcement motivations for SH on the SITBI responded faster to the target on SH trials (rs = −0.316, p = 0.027). SH participants with higher DASS-21 depression subscale scores had significantly more premature responses in SH trials (*rs* = 0.31, *p* = 0.032). Participants in the SH group whose SH-related mental imagery generated more positive emotions (on PANAS score embedded in the SHII) made more correct responses on SH trials (rs = 0.348, p = 0.017). See Supplementary Figure S4.

#### Associations between Incentive Delay Task performance and appraisal of SH stimuli

Ratings for SH stimuli in the three groups are reported in Supplementary Table S1. In the SH group, those who rated SH stimuli as more pleasant and salient (total rating score) had a greater number of correct responses on SH trials (pleasant *rs* = 0.29, *p* = 0.032; total rating score *rs* = 0.29, *p* = 0.034). Distressing and unpleasant ratings were not associated with performance in the SH group. In the NA group, those rating SH stimuli as more unpleasant, distressing and salient had shorter mean RTs on SH trials (unpleasant *rs* = −0.32, *p* = 0.015; distressing *rs* = −0.27, *p* = 0.049; total rating score *rs* = −0.31, *p* = 0.021). There were no significant associations between SH stimuli ratings and IDT performance in the HC group (*p* > 0.05).

### Study 2

Seventy-five SH and 70 HC participants initially completed the Trier Social Stress Test and IDT task. After removal of outliers (10 SH, 7 HC) (see Supplementary Material), a final sample of 65 SH and 63 HC participants was left. Demographic characteristics and scores on self-report questionnaires are reported in Table 2 and 3 respectively. Clinical characteristics are reported in Supplementary Table S4.

**Table 2.**
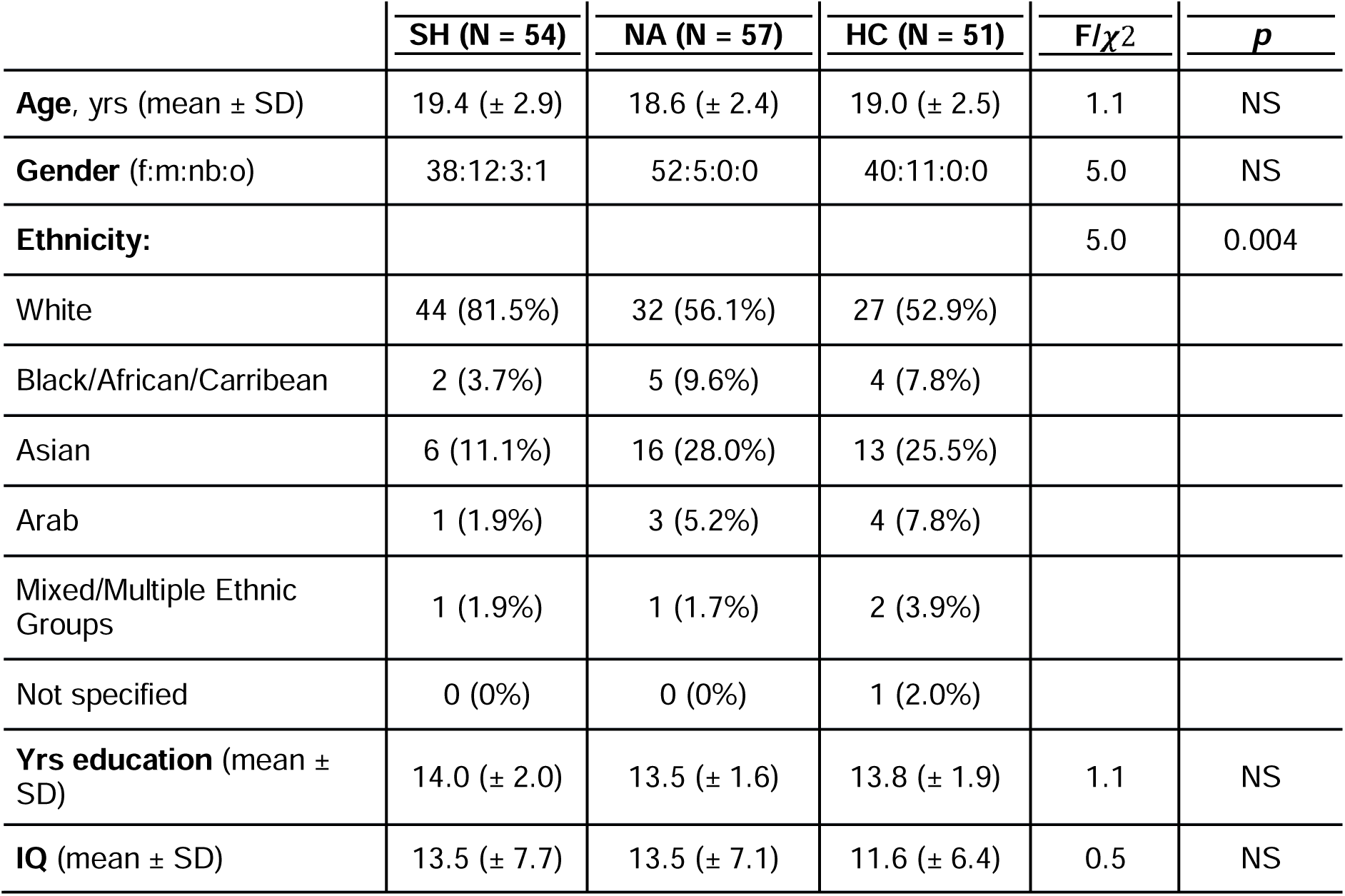
Demographic characteristics of the Study 1 sample. IQ was calculated based on National Adult Reading Test (NART) score.*White vs non-White.

**Table 3.** Demographic characteristics for sample in Study 2. IQ was calculated based on National Adult Reading Test (NART) scores.

| | SH (N = 80) | HC (N = 76) | F/$\chi^2$ | *p* |
| --- | --- | --- | --- | --- |
| **Age, yrs** (mean ± SD) | 18.2 (± 2.4) | 20.4 (± 2.6) | -5.58 | <0.001 |
| **Gender** (f:m:nb) | 71:5:4 | 56:20:0 | 14.68 | <0.001 |
| **Ethnicity:** |  |  |  | NS |
| White | 41 (51%) | 31 |  |  |
| Black/African/Carribean | 8 (10%) | 7 |  |  |
| Asian | 24 (30%) | 34 |  |  |
| Arab | 0 (0%) | 2 |  |  |
| Mixed/Multiple Ethnic Groups | 4 (5%) | 2 |  |  |
| Not specified | 3 (4%) | 0 |  |  |
| **Yrs education** (mean ± SD) | 12.7 (± 5.8) | 15.3 (± 5.6) | -2.81 | 0.006 |
| **IQ** (mean ± SD) | 10.7 (± 8.6) | 11.7 (± 5.6) | -0.84 | NS |

#### Trier Social Stress Test (TSST)

There was a significant main effect of time on negative affect (PANAS scores) (F(1, 236) = 53.69, *p* < 0.001, *ηp2* = 0.19), where negative affect scores were significantly higher following the TSST than before across both SH and HC groups (*p* < 0.0001). There was also a significant main effect of group on negative affect (F(1, 236) = 111.23, *p* < 0.001, *ηp2* = 0.32) and positive affect (PANAS scores) (F(1, 236) = 29.96, *p* < 0.001, *ηp2* = 0.11), where SH participants had significantly higher negative affect scores and lower positive affect scores than HC participants both before and after the TSST (*p* < 0.0001)(Figure 2).

#### Incentive Delay Task performance

There was a significant main effect of reward type on the number of correct responses (F(1, 810) = 47.65, *p* < 0.001, *ηp2* = <0.01), with significantly greater number of correct responses for the money reward than the SH or social rewards across both groups (*p* < 0.0001)(Figure 3). All other main effects and interaction effects were non-significant (*p* > 0.05).

There was a significant main effect of group on RT (F(1, 786) = 8.68, *p* < 0.05, *ηp2* = 0.01), where those in the SH group had significantly faster RTs than the HC group, regardless of the reward type and condition (*p* < 0.05)(Figure 4). All other main effects and interaction effects were non-significant (*p* > 0.05).

There was a significant main effect of condition on the number of premature responses (F(1, 375) = 19.43, *p* < 0.001, *ηp2* = 0.02), with a significantly higher number of premature responses in ‘win’ trials compared with neutral trials across both groups (*p* < 0.0001)(Figure 5). There was a significant main effect of reward type on the number of premature responses (F(2, 461) = 11.93, *p* < 0.001, *ηp2* = 0.03), where participants across both groups had a significantly greater number of premature responses for the money rewards than the SH and social rewards (*p* < 0.05). There was also a significant reward type x condition interaction (F(2, 242) = 6.27, *p* < 0.05, *ηp2* = 0.02), where there were a significantly greater number of premature responses in ‘win’ trials compared with neutral trials for the SH and money rewards across both groups (*p* < 0.05), but not for ‘win’ (positive) social trials compared with neutrals social trials. All other main effects and interaction effects were non-significant (*p* > 0.05).

There was no significant difference in the total amount of money won between the SH and HC groups (*p* > 0.05)(Supplementary Materials).

#### Association between Incentive Delay Task performance and affect change after TSST

There was no significant association between any of the IDT outcome measures and change in positive or negative affect (PANAS scores after TSST - PANAS scores before TSST) in either the SH or HC group (*p* > 0.05).

#### Associations between IDT performance and self-report questionnaires

Total CEQ-SH score was significantly associated with premature responding on SH trials, where a higher urgency to engage in SH was significantly associated with fewer premature responses in the SH group (*rs* = −0.29, *p* = 0.04). All other associations were non-significant (p > 0.05) (Supplementary Figure S5).

## Discussion

Contrary to our hypothesis, young people with SH behaviour did not show a motivational bias towards salient SH-related stimuli on a novel behavioural Incentive Delay Task compared to controls. Moreover, we did not find any biases in behavioural measures of reward anticipation for monetary or social rewards in individuals with SH compared to controls. Negative affect induction via a social stress task was effective at inducing negative affect, but did not modulate the response to either SH-related cues or natural rewards. When exploring SH characteristics, those with fewer years of-, a greater urge to- and a higher automatic positive reinforcement motivation for SH, as well as those with higher positive affect following SH-related mental imagery, made more correct, faster and fewer premature responses on reward trials in the SH condition of the IDT. However, these associations were not replicated across our two studies.

Our results suggest that SH-related stimuli do not acquire an incentive value in young people with a history of SH. However, it is possible that SH stimuli may have triggered bi-directional biases at the individual-level in the SH group, in the form of both approach and avoidance behaviours, thus minimizing any group-level directionality of bias. This would be consistent with ambivalence models reported in substance use^52–54^, other emerging evidence in SH^45,55^ and in those with suicidal ideation^56^. Results from an attentional Dot Probe task also conducted in Study 1, support an avoidance of SH-related cues in the same SH group^55^. As reported in this latter study^55^, members of our YPRG described having both a desire to look at the SH images and to avoid them, and suggested that this may be influenced by the motivation to stop self-harming in keeping with previous literature^57^. It is unclear if and how this is associated with individual differences in SH characteristics such as duration of SH history, given that a shorter duration of SH was associated with faster responses on SH trials in the Incentive Delay Task, but also greater attentional avoidance of SH cues in the Dot Probe Task^55^ in Study 1.

Exploratory analyses tentatively point to other factors that may enhance or contribute towards motivation for self-harm. Specifically, young people who more strongly endorsed self-harming for automatic positive reinforcement, for example “to feel something” or increase positive affect, and rated the SH stimuli as more pleasant had faster and more correct responses on SH trials in Study 1. This finding is consistent with greater attention towards SH cues on the Dot Probe task in the same study^55^. Additionally, young people whose mental imagery of self-harm generated more positive emotions made more correct responses to SH trials. Based on these exploratory findings, future work could investigate if acquisition of an incentive bias towards SH-related stimuli might be present in a sub-group of young people for whom positive reinforcement drives the behaviour, and if emotional response to SH-related imagery may capture this mechanism^12^.

We found a significant correlation between negative affect DASS-21 scores and faster responding to SH images in the SH but not the NA group in Study 1. However, our NA induction in Study 2 refutes the hypothesis that negative affect would elicit sensitisation to SH cues in individuals with SH. This is contrary to findings in reported in addiction^58–60^. A possible explanation could be that TSST-related effects on affect may have been too brief to influence IDT performance, as subjective stress levels have been shown to decrease significantly from the beginning of the arithmetic part of the online TSST to the end^51^. Whilst we found a significant increase in PANAS negative affect after the TSST, this may not have been sufficient to effectively influence IDT task performance, given the difference between our procedure and past studies, including the number of judges and length per section^49,50^. Moreover, TSST effects may be mediated by stress hormones such as cortisol, which in turn modulate reward processing, and stress-response dysfunction has been associated with SH^48,49^. Alternatively, NA may drive SH behaviour via other mechanisms than modulating approach towards SH cues. Previous findings showed that acute stress does not impact responses to rewarding stimuli in patients with Borderline Personality Disorder^61^ and Major Depressive Disorder^62^, which commonly co-occur with SH^2^.

Another explanation for the inability of our NA induction to elicit sensitization to SH cues may be due to heterogeneity in our SH samples in terms of SH characteristics. For example, SH frequency spanned from only once to greater than 400 times in a lifetime (see Table 1). This may explain why some individual differences in specific factors including duration of SH were tentatively associated with IDT performance in the absence of significant between-group differences, but equally why these associations don’t always replicate across our two studies. Future studies should clarify whether SH participants can be clustered based on variables such as SH frequency and whether such clusters may be associated with unique cognitive mechanisms underpinning SH behaviour.

Although previous studies using fMRI have shown reduced neural activity during reward anticipation in SH participants compared with HCs, our findings do not support the presence of biases in processing monetary rewards in young people with SH^17^. Rather than showing reduced anticipation to monetary reward, the SH group performed with better accuracy across all trials and won more money than the control groups in Study 1. It is possible that the feedback “try to be faster” after incorrect responses was perceived as particularly stressful by this population, and motivated them to do better, as reported by our YPRG members. This may have skewed results towards maximised performance across all reward type trials. Interestingly, our NA group in Study 1 also appeared as sensitive to monetary reward as the HC group despite depressive symptoms. This is in line with previous monetary IDT findings showing that low mood per se is not associated with reduced reward anticipation but anhedonia is^63^. Of note, medication use influenced IDT performance, but did not affect our results (Supplementary Tables S6-S8).

### Limitations

A possible limitation of our study might be that as participants received a tangible reward (money) only in the monetary condition, this could have masked more subtle motivational biases towards image-based cues for SH and social rewards. Further, our SH images may not have been salient enough, for example they did not show blood or wounds, while seeing blood during SH is thought to contribute to its reinforcing nature^64^. More ecologically-valid stimuli, such as social media ‘likes’^21^, reactions to text messages or interpersonal situations could have been used in the IDT social condition. It is also possible that simple behavioural markers as our modified IDT are unable to detect reward system biases underpinning SH behaviour. Finally, we did not ask participants about their motivation to stop self-harming or engagement in therapy, which may impact on response to SH-related stimuli; and we did not collect data on SH-related mental imagery in Study 2. Our YPRG also suggested that our sample may have been biased towards individuals trying to stop self-harming, as they would be more likely to take part in research.

### Conclusion

Overall, young people who SH did not show incentive sensitisation to SH stimuli, nor biases in processing social or monetary rewards. This also remained true also following a NA induction, strengthening the phenomenological validity of our findings. As such, from a translational perspective, anticipation of SH-related cues or rewards do not appear to have potential as markers for targeting SH interventions. However, we found an indication that motivational biases may be present in a sub-group of young people with SH. Future studies should replicate our findings either by recruiting large samples powered to robustly capture individual differences with small effect size or by focusing on those who describe their SH as ‘addictive’ and perceive SH-related cues, including mental imagery, as motivating. Furthermore, understanding how ‘addictive’ properties of SH behaviour may relate to greater suicide risk may also help with the development of targeted preventative strategies for SH. The addition of physiological measures such as salivary cortisol may also better elucidate SH profiles for the purpose of risk, trajectory and intervention stratification.

## Data Availability

All materials and data are reported on Open Science Framework (https://osf.io/8kcg3/).

## Acknowledgements

We thank the Young Person’s Research Group: Catherine Thomas, Caroline Crandell, Maya Kamall, Martin Sinclair, Katy McSweeny, Tabitha A B Podger and Abby Martin for their valuable insight and work on this research. We would also like to thank members of the Department of Psychiatry at Imperial College London for acting as judges during the TSST.

