## Supplementary Materials for "Reward processing in young people with self-harm behaviour"

**Methods**

The findings reported here were part of the broader project “iMAGine: motivational abnormalities guiding self-harm and binge-purge behaviour” , see imagine study.org.

*Recruitment*

Participants were recruited using social media advertisements on Instagram and Facebook and reimbursed with a £50 voucher for completing the testing. Young people expressed completed an online screening form on Qualtrics to express their interest in the study, which consisted of questions on age, gender, history of self-harm, the DASS-21, the Cannabis Use Disorder Test-Revised (CUDIT-R) and the Alcohol Use Disorder Identification Test (AUDIT). Participants were then invited to a five minute telephone screening if assessed as eligible based on the online screening form. During the telephone screening, eligibility crtieria were confirmed verbally by participants, and information on methods of self-harm was collected in order to prepare the self-harm computer tasks prior to the testing. If participants were deemed eligible during the telephone screening, they were then invited to the video-call screening on Microsoft Teams. During this video screening, participants first consented to participate in this study, after which they completed the DASS-21, MINI, smoking assessment, McLean Screening Instruement for Borderline Personality Disorder (MSI-BPD) and the NART. Participants in the SH group also completed the NSSI section of the Self-Injurious Thoughts and Behaviour Interview (SITBI), Alexian Brothers Urge to Self-harm scale (ABUSI) and the Self-harm Images Interview (SHII).

Experimental procedure

Testing immediately followed the video-call screening and was c arried out on Microsoft Teams. Testing consisted of two 1.5 hour sessions. Participants were asked to keep their cameras on where possible during the testing to improve compliance. As testing was not lab-based, any interruptions, distractions or technical difficulties during testing were recorded and these participants were excluded from analyses where appropriate.

Participants answered self-report questionnaires via Qualtrics links shared by the experimenter over MS Teams. Clinical characteristics were assessed via the MINI^36^ and the McLean Screening Instrument for Borderline Personality Disorder (MSI-BPD)^37^. Questionnaires relating to SH included the Self-Injurious Thoughts and Behaviour Interview (SITBI)^38^, the Self-Harm Imagery Interview (SHII)^39^ (Study 1 only), the Alexian Brothers Urge to Self-Injure Scale (ABUSI)^40^ and the Craving Experiences Questionnaire adapted for SH (CEQ-SH)^13^ (Study 2 only). State depression, anxiety and stress was assessed using the DASS-21^41^. The Positive and Negative Affect Schedule (PANAS) was administered before and after the Trier Social Stress Test in Study 2. In addition to the tasks described here, participants also completed additional tasks that are part of additional publications.

Following the completion of these questionnaires, participant completed the Incentive Delay Task (IDT) in both studies. In study 2 however, participants first underwent the Trier Social Stress Test (TSST) followed by the IDT. The TSST always preceeded the IDT in Study 2.

Self-report questionnaires

The **Depression, Anxiety and Stress Scale** (DASS-21)^36^ measures state depression, anxiety and stress, and has acceptable to excellent internal consistency in clinical and non-clinical samples^36^. The depression scale measures symptoms associated with dysphoric mood e.g. worthlessness, the anxiety scale measures symptoms associated with physiological arousal such as trembling, and the stress scale measures symptoms of tension and reactivity to stressful events.

habit proneness. Participants with restrictive eating disorders were excluded from part two of the scale

The **Alcohol use disorders Identification Test** (AUDIT)^65^ screens for hazardous or harmful drinking behaviour, measuring alcohol misuse and possible dependence.

The **Cannabis Use Disorder Identification Test Revised** (CUDIT-R)^66^ screens for problem cannaibis use, with excellent internal consistency.

The **smoking assessment** involved asking participants about their smoking and vaping:

1. Do you smoke? (Yes/No/Occassionally)
2. If yes, how many cigarettes on an average day/week/month/year?
3. Do you vape? (Yes/No/Occassionally)
4. If yes, how many times do you vape on an average day/week/month/year?

The **MINI International Neuropsychiatric Interview (MINI)**^37^ is a short structured diagnostic interview carried out by trained researchers, exploring 17 disorders according to the DSM-IV diagnostic criteria.

The **McLean Screening Instrument for Borderline Personality Disorder (MSI-BPD)**^38^ screens for Borderline Personality Disorder (BPD) with all ten items based on the DSM-IV diagnostic criteria for BPD. Two items are based on the ninth DMS-IV criteria for paranoia/dissociation and each of the remaining eight items are based on the remaining eight DSM-IV criteria.

**Medication Use**: participants were asked ‘are you taking any regular medication for anxiety, depression, ADHD etc? If so, please specify?’

The **National Adult Reading Test** (NART)^67^ is a widely-used proxy for IQ in clinical research.

The **Self-Injurious Thoughts and Behaviours Interview** (SITBI)^39^ assesses various aspects of self-harm behaviour including frequency, methods and function, with excellent inter-rater reliability and test-retest reliability. Self-harm methods were collected by asking participants how they self-harm. Specific methods were not listed to participants in order to reduce potential distress or increased use of such methods as a result of completing the questionnaire, as advised by the YPRG. One question on recency of self-harm behaviour was also added: ‘when was the last time you self-harmed?’. Only the non-suicidal self-injury section of this interview was used, replacing the term ‘non-suicidal self-injury’ with ‘self-harm’ given that our definition includes all self-harm behaviour regardless of intent.

The **Alexian Brothers Urge to Self-Injure Scale** (ABUSI)^41^ assesses the intensity of the urge to self-harm over the past week, capturing emotional and cognitive aspects of how motivated someone is to engage in self-harm.

The **Self-harm Images Interview** (SHII): Self-harm imagery was assessed using a 10-item semi-structured interview, measuring image content, appraisal of the image, and emotions experienced after the image. The scale was adapted from the Mental Imagery Interview^68^, as already used in a previous study^40^.

Incentive Delay Task (IDT)

The stimuli used for used for each of the three conditions (SH, Social and Money) in this task are described below.

**SH condition stimuli:** SH stimuli were images of SH acts (Figure 4). ‘No self-harm’ stimuli were close-up images of hands holding neutral objects (Figure 4). All stimuli in the SH condition were created by the research team. Before testing, the ten SH stimuli that were identical across tasks and ‘no self-harm’ stimuli were rated by nine university students on a scale of 0 (not at all) to 10 (extremely) for calming, exciting, pleasant, unpleasant and distressing emotions (Supplementary Table S1). Following this, five ‘no self-harm’ stimuli were removed as they could be interpreted as positive by participants and replaced by neutral images similar to those already included.

**Monetary condition stimuli:** Win money stimuli consisted of images of different containers with the same amount of money, and no money stimuli were the same containers without money, with all images created by the research team.

**Social condition stimuli:** To improve ecological validity of the task social stimuli were images of people in naturalistic settings, compared to the individual faces against a plain background commonly used in Social Incentive Delay tasks^69,70^. Twenty positive social and twenty neutral social stimuli were chosen by the Young Person’s Research Group (YPRG) using an online search and were rated from 0 (not at all) to 10 (extremely) for connected, exciting, upsetting and boring emotions by nine university students (Supplementary Table S2). Neutral stimuli with scores above five for calming, exciting and upsetting were removed and positive stimuli with scores above 5 for upsetting and boring were removed. These removed images were replaced by new images chosen by the same YPRG.

See Figure S1 for images of the stimuli used in the Incentive Delay Task.

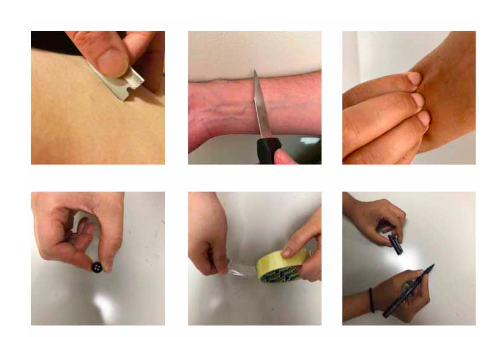

A)

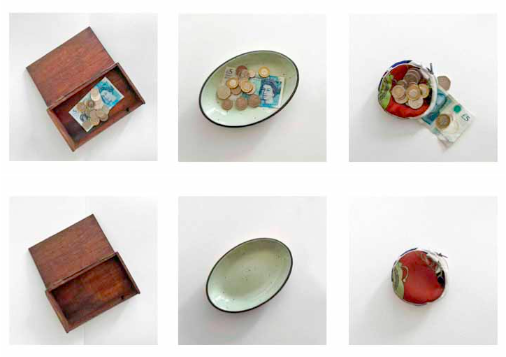

B)

**Figure S1.** Stimuli used in the Incentive Delay Task, showing self-harm stimuli (A top), no self-harm stimuli (A bottom), win money stimuli (B top) and no money stimuli (B bottom).

Trier Social Stress Test (TSST)

The protocol for the TSST was based on an online version of the task developed by a recent study^51^. The TSST consisted of 3 sections: a 5-minute speech preparation period, a 5-minute speech presentation, and a 5-minute arithmetic task. The speech and arithmetic task are completed in front of 2 judges. Before starting the task, the experimenter informed the participant that they would leave the call for the task and then rejoin once the task had finished. The experimenter then called the 2 judges onto the teams call, and then left teams call once the two judges had joined. The 2 judges appeared on-screen as ‘Judge 1’ and ‘Judge 2’, with their camera and audio turned on, and were logged in using separate accounts. The 2 judges informed the participant that they would have 5 minutes to prepare a speech where they would introduce themselves and list several good and not so good things about themselves (based on a recent study^64^). The 2 judges then turned off their video to mark the start of the 5-min speech preparation time. After 4 minutes of the speech preparation had passed, the female judge announced “you have one minute left until you have to give your speech”. At the end of the 5 minutes of speech preparation, the judges turned their cameras on and the female judge announced “make sure you can see both of us and please make sure your phone is turned off during the speech”. The male judge then announced “your speech should be 5 minutes long, and if you finish early, you will be told to continue. You may start your speech …(pause)... now”. If a participant ended their speech before 5 minutes had past, they were told to continue until the end of the 5 minutes. Once five minutes of the speech were up, the male judge interrupted and said “your time is now up” and then moved onto the third and final 5-min arithmetic part of the task. The male judge then announced “for this final part of the task, you must subtract the number 13 from the number 938. If you make a mistake, you will be asked to start over.” The participant then began to count down from 938. If the participant made a mistake, the female judge interrupted and told them to start again. If they were going too slowly, the male judge interrupted and told them to start again, making sure they were subtracting as quickly and accurately as possible. If the participant made many errors in a row, the female judge interrupted and told them to subtract from 938 by the number 7, and if they then continued to make errors, by the number 3. If the participant did not make any mistakes whilst subtracting by 13, the female judge interrupted and told them to subtract by 17. At the end of the 5-minute arithmetic period, the male judge interrupted and said “your time is now up. Thank you for completing this task. You will now be passed back to the experimenter”. The two judges then left the call and the experimenter rejoined the call. Although not always possible, an effort was made to ensure that one of the judges was female and one was male, as in the protocol used by recent studies^51,64^.

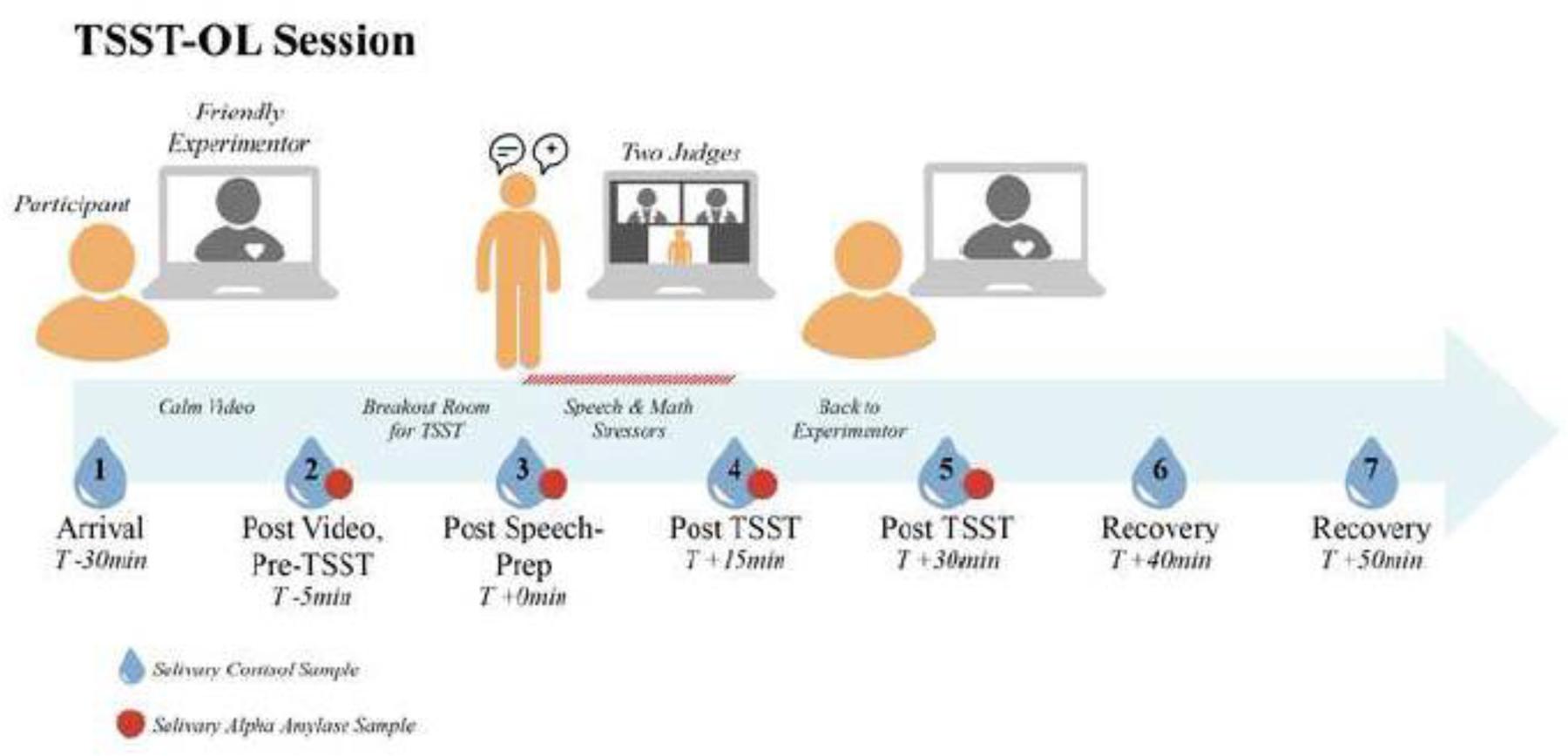

Figure S2: Trier Social Stress Test (TSST), adapted from a previous study^51^.

*Statistical analysis*

Power analysis

A power analysis using a t-test power calculation was conducted based on data from a study^17^ that compared RTs of SH adolescents with HCs using an fMRI Monetary Incentive Delay task. This indicated that N=48 participants were needed per group for an effect size of d=0.580 at 80% power. A loss of N=5 was estimated per group to account for poor task performance and attrition, setting the target for recruitment at N=53 participants per group. In Study 2 participants were over-recruited to account for power calculations for another experimental task (reported elsewhere).

Outlier removal

Outliers were removed based on whether there premature response, correct response or RT scores on the IDT were 1.5 x Interquartile Range (IQR) below or above the lower or upper quartile respectively. For Study 1, 9 outliers were removed in total (2 HC, 2 NA and 5 SH). 1 HC participant was removed due to missing data, 1 HC, 1 NA and 3 SH participants due to task-irrelevant distractions, 1 NA participant due to outlier premature response scores, 1 SH participant due to technical failures and 1 SH participant as they reported using items shown in neutral SH trials to self-harm. This resulted in a final sample of 49 SH, 54 NA and 49 HC valid data for the IDT. These same excluded participants were also excluded from the SH stimuli rating analysis, with 1 additional NA participant removed due to missing data. For Study 2, 17 outliers were removed in total (10 SH and 7 HC). 3 SH and 5 HC participants were removed due to outlier RTs, 1 SH and 2 HC participants due to outlier premature responses and 6 SH participants due to outlier correct responses on the IDT. This resulted in a final sample of xx SH and xx HC valid data for the IDT.

**Results**

*Study 1*

*Sample*

|  | **SH (N = 54)** | **NA (N = 57)** | **HC (N = 51)** |
| --- | --- | --- | --- |
| **DASS-21**, mean (± SD): |  |  |  |
| Depression | 20.7 (± 10.6) | 18.1 (± 9.1) | 2.3 (± 2.1) |
| Anxiety | 15.0 (± 9.4) | 14.2 (± 8.6) | 1.6 (± 1.8) |
| Stress | 21.7 (± 9.6) | 20.1 (± 9.3) | 4.9 (± 3.7) |
| Total | 19.1 (± 8.5) | 17.5 (± 7.6) | 2.9 (± 1.7) |
| **AUDIT** (mean ± SD) | 6.6 (± 7.3) | 3.5 (± 4.0) | 3.1 (± 3.6) |
| **CUDIT-R** (mean ± SD) | 1.8 (± 3.3) | 0.6 (± 2.6) | 0.24 (± 0.9) |
| **Nicotine** cigarettes, yes:occasionally:no,  % (N) | 9(15):17(9):74(40) | 0:0:100(57) | 0:6(3):94(48) |
| **Nicotine** vaping, yes:occasionally:no,  % (N) | 4(2):4(2):92(50) | 0:2(1):98(56) | 0:2(1);98(50) |
| **MSI-BPD** (mean ± SD) | 6.6 (± 2.5) | 0.6 (± 1.9) | 0.4 (± 1.0) |
| MSI-BPD criteria met, yes:no, % (N) | 46(25): 54(26) | 12(7):88(50) | 0:100(50) |
| **MINI Neuropsychiatric Interview** % (N) |  |  |  |
| Major Depression (current:recurrent) | 48(26):28(15) | 35(20):23(13) |  |
| Dysthymia current | 3 | 5 |  |
| Suicide risk (none:low:moderate:high) | 4(2):48(26):30(16):19(10) | 88(50):12(7):0:0 |  |
| Hypomanic episode (current:past) | 0:15(8) | 0:9(5) |  |
| Manic episode (current:past) | 0:11(6) | 2(1):9(5) |  |
| Panic disorder (current:past) | 6(3):39(21) | 7(4):23(13) |  |
| Panic disorder limited symptoms lifetime | 28(14) | 14(8) |  |
| Agoraphobia current | 46(25) | 39(22) |  |
| Social phobia current | 26(14) | 18(10) |  |
| Obsessive compulsive disorder current | 19(10) | 5(3) |  |
| Post-traumatic stress disorder current | 17(9) | 9(5) |  |
| Alcohol use current (abuse:dependence) | 2(1):22(12) | 0:2(1) |  |
| Substance use current (abuse:dependence) | 0:9(5) | 0:0 |  |
| Mood disorder with psychotic features (current:past) | 4(2):7(4) | 0 |  |
| Psychotic disorder (current:past) | 0:0 | 0:0 |  |
| Bullimia Nervosa current | 7(4) | 2(1) |  |
| Anorexia Nervosa current | 2(1) | 0 |  |
| Anorexia binge-purge type current | 0 | 0 |  |
| Generalised anxiety current | 41(22) | 51(29) |  |

**Table S1.** Clinical and affective measures of the sample in Study 1.

|  | **SH (N = 54)** | **NA (N = 57)** | **HC (N = 51)** |
| --- | --- | --- | --- |
| **Current psychoactive medication use** % (N): |  |  |  |
| Reuptake inhibitor (SERT) | 33 (18) | 9 (5) | 0 |
| Receptor antagonist (D2, 5-HT2), Reuptake inhibitor (NET) | 6 (3) | 0 | 0 |
| Receptor agonist (MEL1, MEL2) | 6 (3) | 0 | 0 |
| Reuptake inhibitor (SERT, NET) | 4 (2) | 0 | 0 |
| Reuptake inhibitor (DAT, NET) Releaser (DA, NE) | 4 (2) | 0 | 0 |
| Reuptake inhibitor (SERT), Receptor agonist (5-HT1A) | 2 (1) | 0 | 0 |
| Receptor antagonist (5-HT2) |  |  |  |
| Benzodiazepine receptor agonist | 2 (1) | 0 | 0 |
| Alpha-2 delta calcium blocker | 2 (1) | 0 | 0 |
| Receptor antagonist (NE alpha-2, 5-HT2, 5-HT3) | 2 (1) | 0 | 0 |
| Receptor partial agonist (D2, 5-HT1A), Receptor antagonist (5-HT2A) | 2 (1) | 0 | 0 |
| Receptor antagonist (H1, D2) | 2 (1) | 0 | 0 |
| Mode of action unknown (Valproate, Levetiracetam) | 2 (1) | 0 | 2 (1) |
| % of participants using psychoactive medication | 64.8% | 9% |  |

**Table S2.** Medication use for Study 1 sample.

IDT performance

Total money won

The SH group won an average of £6.92 (± £1.46), the NA group £6.19 (± £1.68), and HC group £6.10 (± £1.70) out of a maximum of £10. A significant main effect of group for money won (F(2, 149) = 10.028, p = 0.025, ηp2 = 0.048) showed that the SH group won significantly more money than the HC group (p = 0.041), but not the NA group (p = 0.073).

Associations between IDT performance and self-report questionnaires

A)
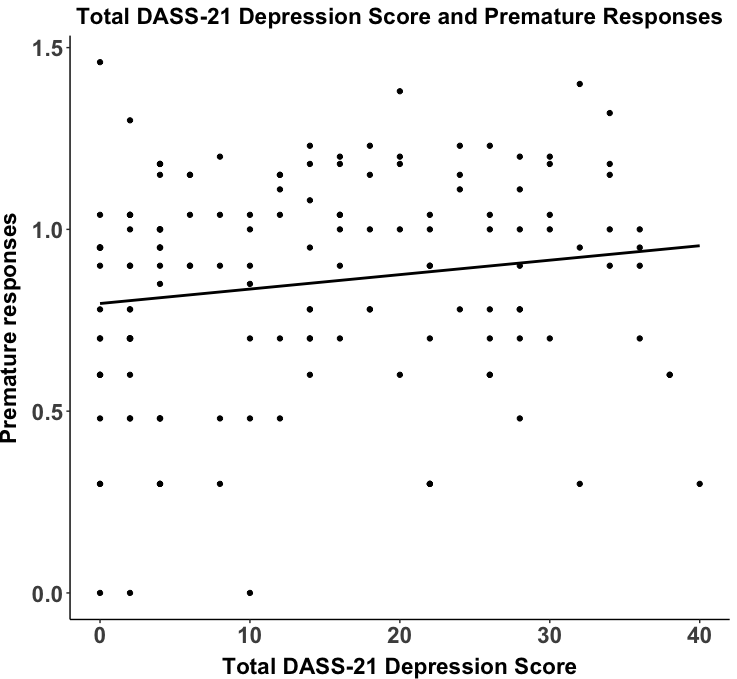

B)
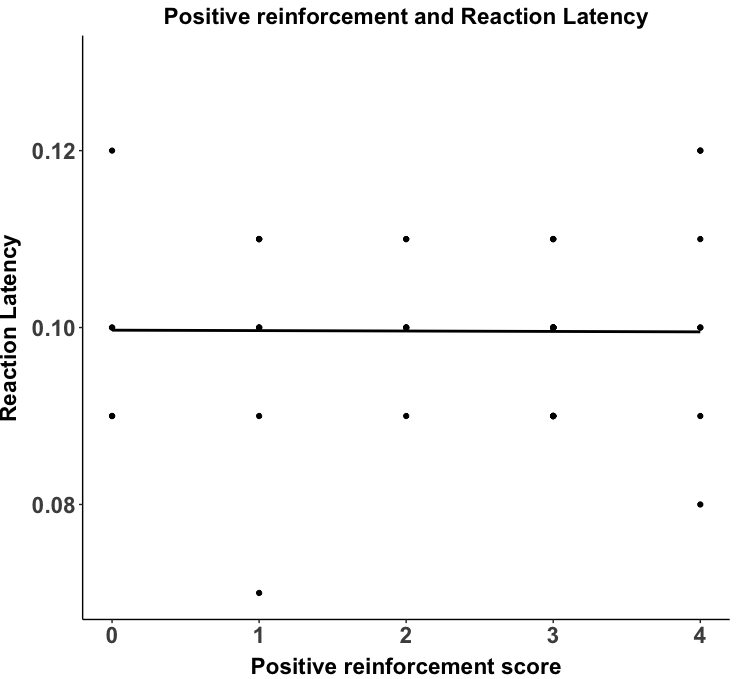

C)
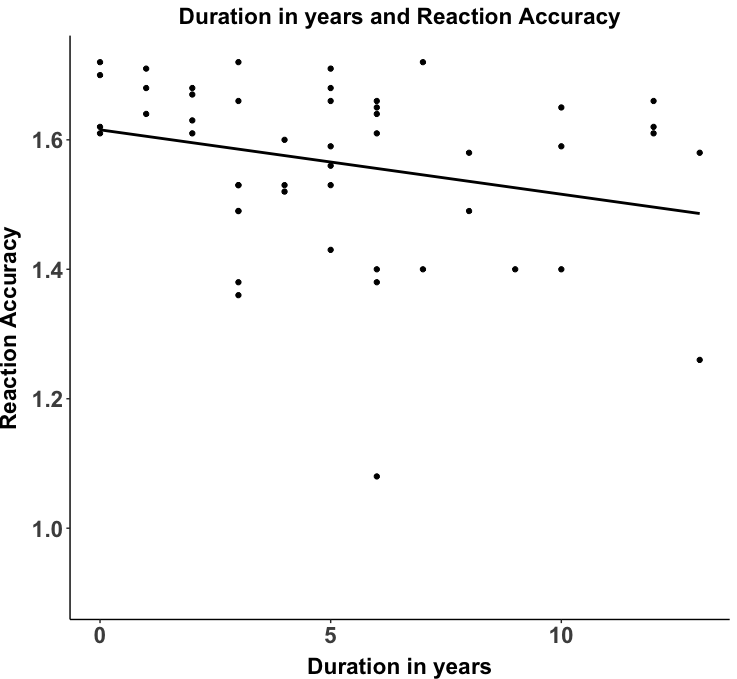

D)
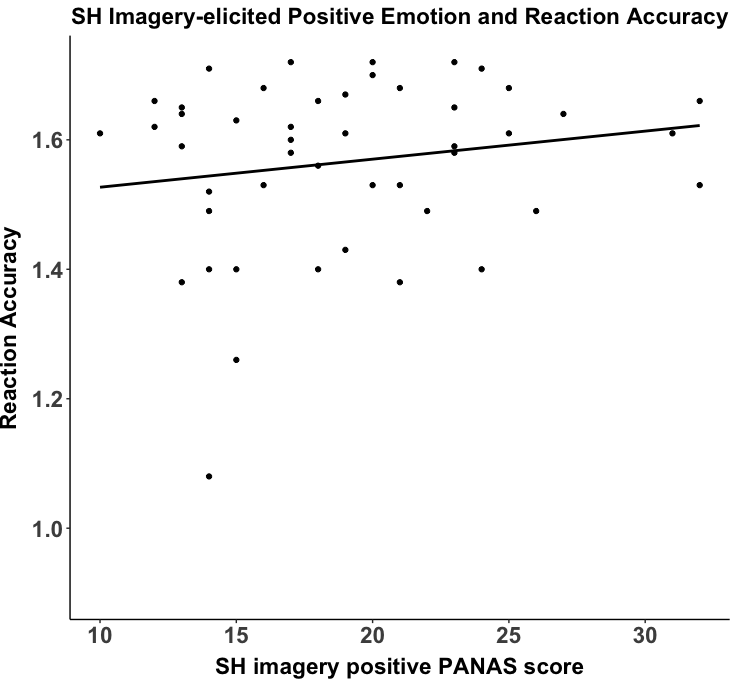

**Figure S4.** Associations between each of the self-harm characteristics and Incentive Delay Task scores in the reward condition of the SH trials for Study 1.

Appraisal of SH stimuli

| Rating | Self-harm | Negative Affect | Healthy Control |
| --- | --- | --- | --- |
| Calming, median  Mean (SD) | 0.0 (0.0)  0.1 (0.4) | 0.0 (0.0)  0.0 (0.1) | 0.2 (1.3)  0.9 (1.3) |
| Exciting, median  Mean (SD) | 0.0 (0.0)  0.1 (0.4) | 0.0 (0.0)  0.0 (0.1) | 0.6 (2.4)  1.3 (1.6) |
| Pleasant, median | 0.0 (0.0) | 0.0 (0.0) | 0.1 (1.1) |
| Mean (SD) | 0.1 (0.3) | 0.0 (0.1) | 0.6 (0.9) |
| Unpleasant, median | 6.2 (2.9) | 6.5 (4.5) | 5.3 (3.0) |
| Mean (SD) | 5.9 (2.3) | 6.0 (2.6) | 5.2 (2.3) |
| Distressing, median  Mean (SD) | 5.8 (2.8)  5.7 (2.3) | 6.1 (4.4)  5.8 (2.8) | 5.4 (3.6)  5.2 (2.5) |
| Total, median  Mean (SD) | 250.0 (127.5)  243.1 (96.1) | 263.5 (167.7)  240.8 (108.9) | 289.0 (180.0)  268.6 (108.4) |

**Table S3.** Mean ± standard deviation and median ratings for self-harm stimuli in the Incentive Delay Task for Self-harm, Negative Affect and Healthy Control groups. Ratings were on a Likert scale from 0 (not at all) to 10 (extremely).

*Study 2*

*Sample*

|  | **SH (N = 80)** | **HC (N = 76)** |
| --- | --- | --- |
| **DASS-21**, mean (± SD): |  |  |
| Depression | 22.5 (± 10.9) | 2.1 (± 2.6) |
| Anxiety | 19.0 (± 10.5) | 1.8 (± 2.3) |
| Stress | 23.7 (± 9.3) | 4.3 (± 3.5) |
| Total | 65.3 (± 26.4) | 8.2 (± 6.4) |
| **AUDIT** (mean ± SD) | 5.5 (± 5.9) | 3.8 (± 3.2) |
| **CUDIT-R** (mean ± SD) | 2.0 (± 4.0) | 0.2 (± 0.7) |
| **Nicotine** cigarettes, yes:occasionally:no:unknown, % (N) | 5(4):21(17):71(57):3(2) | 1.3(1):1.3(1):96(73):1.3(1) |
| **Nicotine** vaping, yes:occasionally:no:unknown, % (N) | 2.5(2):20(16):75(60):2.5(2) | 1.3(1):0(0);97.3(74):1.3(1) |
| **MSI-BPD** (mean ± SD) | 6.7 (± 2.4) | 0.3 (± 0.5) |
| MSI-BPD criteria met, yes:no:unknown, % (N) | 57.5(46):30(24):12.5(10) | 0:100(76):0 |
| **MINI Neuropsychiatric Interview** % (N) |  |  |
| Major Depression (current:recurrent) | 51(41):35(28) |  |
| Dysthymia current | 19(15) |  |
| Suicide risk (none:low:moderate:high:unknown) | 40(32):20(16):21(17): 18(14):1(1) |  |
| Hypomanic episode (current:past) | 4(3):20(16) |  |
| Manic episode (current:past) | 8(6):11(9) |  |
| Panic disorder (current:past) | 13(10):44(35) |  |
| Panic disorder limited symptoms lifetime | 13(10) |  |
| Agoraphobia current | 43(34) |  |
| Social phobia current | 43(34) |  |
| Obsessive compulsive disorder current | 15(12) |  |
| Post-traumatic stress disorder current | 14(11) |  |
| Alcohol use current (abuse:dependence) | 1(1):10(8) |  |
| Substance use current (abuse:dependence) | 1(1):5(4) |  |
| Mood disorder with psychotic features (current:past) | 0(0):0(0) |  |
| Psychotic disorder (current:past) | 0(0):0(0) |  |
| Bullimia Nervosa current | 6(5) |  |
| Anorexia Nervosa current | 0(0) |  |
| Anorexia binge-purge type current | 1(1) |  |
| Generalised anxiety current | 4(32) |  |

**Table S4.** Clinical and affective measures for Study 2 sample.

|  | **SH (N = 80)** | **HC (N = 76)** |
| --- | --- | --- |
| **Current psychoactive medication use % (N):** |  |  |
| **Reuptake inhibitor (SERT)** | 26.3 (21) | 0 |
| **Receptor antagonist (D2, 5-HT2), Reuptake inhibitor (NET)** | 0 | 0 |
| **Receptor agonist (MEL1, MEL2)** | 0 | 0 |
| **Reuptake inhibitor (SERT, NET)** | 5 (4) | 0 |
| **Reuptake inhibitor (DAT, NET) Releaser (DA, NE)** | 2.5 (2) | 0 |
| **Reuptake inhibitor (SERT), Receptor agonist (5-HT1A)** | 0 | 0 |
| **Receptor antagonist (5-HT2)** |  |  |
| **Benzodiazepine receptor agonist** | 0 | 0 |
| **Alpha-2 delta calcium blocker** | 0 | 0 |
| **Receptor antagonist (NE alpha-2, 5-HT2, 5-HT3)** | 2.5 (2) | 0 |
| **Receptor partial agonist (D2, 5-HT1A), Receptor antagonist (5-HT2A)** | 0 | 0 |
| **Receptor antagonist (H1, D2)** | 2.5 (2) | 0 |
| **Mode of action unknown (Valproate, Levetiracetam)** | 0 | 0 |
| **% of participants using psychoactive medication** | 37.5% | 0 |

**Table S5.** Medication use for or Study 2 sample.

IDT performance

Total money won

There was no significant difference in the total amount of money won between the SH and HC groups (*p* > 0.05)(Supplementary Materials).

Associations between IDT performance and self-report questionnaires

Total CEQ-SH score was significantly associated with premature responding on SH trials, where a higher urgency to engage in SH was significantly associated with fewer premature responses in the SH group (*rs* = -0.29, *p* = 0.04)(Supplementary Figure S5). All other correlations were non-significant (*p* > 0.05).

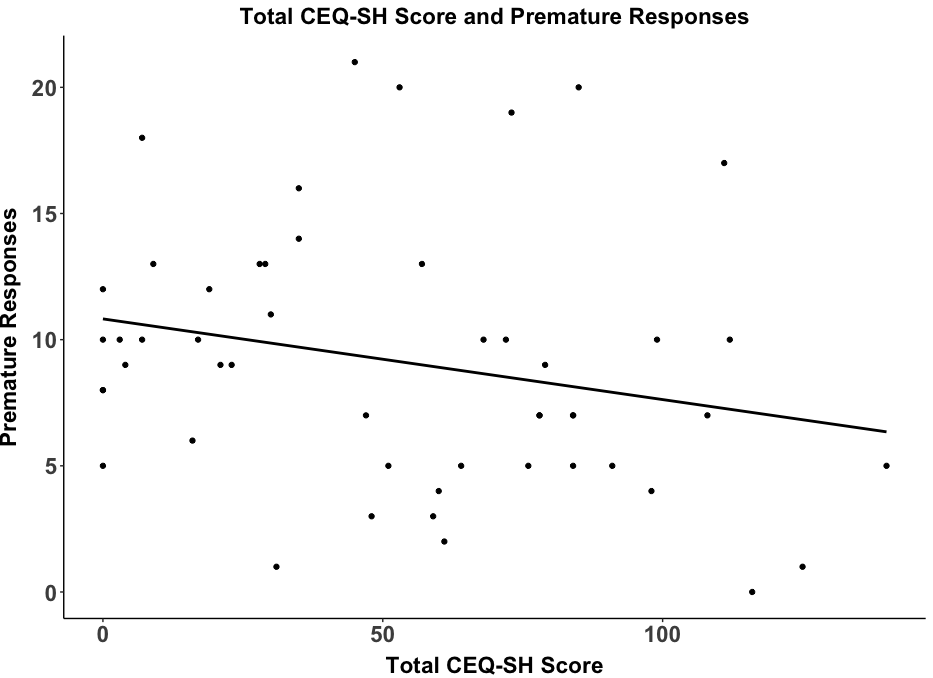

A)

**Figure S5.** Associations between each of the self-harm characteristics and Incentive Delay Task scores in the reward condition of the SH trials for Part 2 of the study.

Medication effects

To control for medication use, an ANCOVA was run using the same variables as in the main model but including medication use (1 = yes, 0 = no) as a covariate. The outcome remained the same when including medication as a covariate.

| **Variable** | **Sum sq** | **Df** | **F** | ***p*** | ***ηp2*** |
| --- | --- | --- | --- | --- | --- |
| Group | <0.01 | 2 | 8.80 | **<0.001** | 0.02 |
| Condition | <0.01 | 2 | 0.99 | 0.373 | <0.01 |
| Reward | <0.01 | 1 | 3.15 | 0.076 | <0.01 |
| Medication | 0.01 | 24 | 4.52 | **<0.001** | 0.11 |
| Group:Condition | <0.01 | 4 | 0.04 | 0.997 | <0.01 |
| Group:Reward | <0.01 | 2 | 0.08 | 0.920 | <0.01 |
| Condition:Reward | <0.01 | 2 | 1.35 | 0.260 | <0.01 |
| Group:Condition:Reward | 0.08 | 4 | 0.47 | 0.760 | <0.01 |

**Table S6. Incentive Delay Task Reaction Latency scores when controlling for medication use. Significant p-values are highlighted in bold.**

| **Variable** | **Sum sq** | **Df** | **F** | ***p*** | ***ηp2*** |
| --- | --- | --- | --- | --- | --- |
| Group | 0.73 | 2 | 24.99 | **<0.001** | 0.05 |
| Condition | 0.49 | 2 | 16.86 | **<0.001** | 0.04 |
| Reward | <0.01 | 1 | 0.05 | 0.819 | <0.01 |
| Medication | 1.92 | 24 | 5.46 | **<0.001** | 0.13 |
| Group:Condition | 0.02 | 4 | 0.27 | 0.896 | <0.01 |
| Group:Reward | 0.01 | 2 | 0.33 | 0.718 | <0.01 |
| Condition:Reward | 0.05 | 2 | 1.59 | 0.205 | <0.01 |
| Group:Condition:Reward | 0.03 | 4 | 0.57 | 0.686 | <0.01 |

**Table S7. Incentive Delay Task Reaction Accuracy scores when controlling for medication use. Significant p-values are highlighted in bold.**

| **Variable** | **Sum sq** | **Df** | **F** | ***p*** | ***ηp2*** |
| --- | --- | --- | --- | --- | --- |
| Group | 2.04 | 2 | 13.59 | **<0.001** | 0.03 |
| Condition | 1.93 | 2 | 12.83 | **<0.001** | 0.03 |
| Reward | 1.58 | 1 | 21.10 | **<0.001** | 0.02 |
| Medication | 7.51 | 24 | 4.17 | **<0.001** | 0.10 |
| Group:Condition | 0.11 | 4 | 0.36 | 0.837 | <0.01 |
| Group:Reward | 0.07 | 2 | 0.45 | 0.637 | <0.01 |
| Condition:Reward | 1.44 | 2 | 9.57 | **<0.001** | 0.02 |
| Group:Condition:Reward | 0.18 | 4 | 0.59 | 0.667 | <0.01 |

**Table S8. Incentive Delay Task Premature Response scores when controlling for medication use. Significant p-values are highlighted in bold.**
